# Features of patients that died for COVID-19 in a Hospital in the south of Mexico: A observational cohort study

**DOI:** 10.1101/2020.09.21.20199117

**Authors:** Jesús Arturo Ruíz-Quiñonez, Crystell Guadalupe Guzmán-Priego, German Alberto Nolasco-Rosales, Carlos Alfonso Tovilla-Zarate, Oscar Israel Flores-Barrientos, Víctor Narváez-Osorio, Guadalupe del Carmen Baeza-Flores, Thelma Beatriz Gonzalez-Castro, Carlos Ramón López-Brito, Carlos Alberto Denis-García, Agustín Pérez-García, Isela Esther Juárez-Rojop

## Abstract

**Background:** Due to the wide spread of SARS-CoV2 around the world, the risk of death in individuals with metabolic comorbidities has dangerously increased. Mexico has a high number of infected individuals and deaths by COVID-19, as well as an important burden of metabolic diseases. However, reports about features of Mexican individuals with COVID-19 are scarce. The aim of this study was to evaluate demographic features, clinical characteristics, and the pharmacological treatment of individuals who died by COVID-19 in the south of Mexico.

**Methods:** We performed an observational study including 185 deceased individuals with confirmed diagnosis of COVID-19. Data were retrieved from medical records. Categorical data was expressed as proportions (%) and numerical data were expressed as mean ± standard deviation. Comorbidities and overlapping symptoms where plotted as Venn diagrams. Drug clusters were plotted as dendrograms.

**Results:** The mean age was 59.53 years. There was a male predominance (60.1%). The mean hospital stay was 4.75 ± 4.43 days. The most frequent symptoms were dyspnea (88.77%), fever (71.42%) and dry cough (64.28%). Present comorbidities were diabetes (60.63%), hypertension (59.57%) and obesity (43.61%). The main drugs used were azithromycin (60.6%), hydroxychloroquine (53.0%) and oseltamivir (27.3%).

**Conclusions:** Mexican individuals who died of COVID-19 had shorter hospital stays, higher frequency of shortness of breath, and higher prevalence of diabetes compared with individuals from other countries. Also, there was a high frequency of off-label use of drugs for their treatment.

## Introduction

On December 31, 2019, the World Health Organization (WHO) was informed of cases of pneumonia of unknown etiology in Wuhan, China. The Chinese authorities identified a new type of coronavirus, which was isolated on January 7, 2020 (1). Coronaviruses are enveloped RNA viruses related to SARS (Severe Acute Respiratory Syndrome) and MERS (Middle East Respiratory Syndrome) coronaviruses. The novel coronavirus was named SARS-CoV2 and COVID-19 is the name of the disease it causes (2, 3). Since it was discovered, SARS-CoV-2 has widely spread around the world; by August 15^th^ 2020, there were 21,037,564 confirmed cases and 755,455 deaths reported (4).

Up to date, there are some reports and reviews about COVID-19 characteristics in individuals from a few populations around the world. These reports agree that the median age of hospitalized individuals is between 47 and 73 years, with a male predominance (60%). Also, the most common symptoms in hospitalized patients are fever (90%), dry cough (60 – 86%), shortness of breath (53 – 80%), fatigue (38%), nausea/vomiting or diarrhea (15 – 39 %), and myalgia (15 – 44%). The most frequent comorbidities observed in hospitalized individuals included hypertension (48 – 57%), diabetes (17 – 34%) and cardiovascular diseases (21 – 28%). Comorbidities are more common in hospitalized individuals (60 – 90%) compared with the overall infected population (25%) (5).

In Mexico, the first case of COVID-19 was detected on February 27, 2020 (6); whereas on August 15^th^, there were 505,751 confirmed cases and 55,293 deaths (4). Researches suggest that an important burden of metabolic diseases such as diabetes (10.3%), hypertension (18.4%) and obesity (36.1%) is present in the Mexican population (7), therefore this population could be highly vulnerable to COVID-19.

Up to today, there are few reports that include epidemiological data to evaluate comorbidities as risk factors in the Mexican population (8, 9). Additionally, we found only one report evaluating gastrointestinal symptoms in Mexican patients with COVID-19 (10).

We believe it is important to evaluate which pharmacological treatment are used. Although there is not directed antiviral therapy recommended against SARS-CoV-2 (11-13), in our sample some off-label drugs were administrated regardless of what has been observed in clinical trials, particularly at the beginning of the pandemic. Therefore, our aim was to evaluate demographic features, clinical characteristics, and pharmacological treatment received by individuals who died by COVID-19 in a third level hospital in the south of Mexico.

## Methods

This was an observational retrospective study performed at the High Specialty Regional Hospital “Dr. Juan Graham Casasús” (HJGC) in Villahermosa, Tabasco, Mexico. We used the medical records of every individual with COVID-19 as the cause of death, who was admitted to the HJGC from April 15th to May 12th, 2020. These individuals had been diagnosed with COVID-19 by epidemiologists following the Mexican Health Secretary’s guidelines; additionally, they tested positive for SARS-CoV-2 by Reverse Transcriptase Polymerase Chain Reaction. The ethical approval was granted by the Research Ethics Committee of the Juárez Autonomous University of Tabasco (103/CIP-DACS/2020) in Mexico.

Three physician doctors retrieved data from clinical files after patients’ death. Data collected included age, gender, clinical symptoms, underlying comorbidities, length of hospital stay, and drugs used as COVID-19 treatment.

We performed a descriptive report. Numerical data was expressed as mean ± standard deviation and proportions (%) for categorical data. Gender, age and comorbidities were compared using the Chi-squared test, with significance of p = 0.05. Comorbidities and overlapping symptoms where plotted as Venn diagrams. Drug clusters were plotted as dendrograms. All data were analyzed using SPSS v.23.

## Results

In this study, the files of 185 deceased individuals were included; these individuals had been diagnosed and treated for COVID-19 between April 15 and May 12, 2020 at the HJGC in Villahermosa Tabasco, Mexico. The length of time in hospital was 4.75 days in means (S.D. 4.43). The distribution of days of hospitalization by gender is shown in Fig 1.

**Fig 1.** Individuals who died of COVID-19 stratified by gender and days of hospitalization.

## Age and Gender

The mean age of the individuals studied was 59.53±12.50 (range 24-86) years. When we stratified them by age, the majority of deaths occurred in individuals 65+ years (36.7%, n=58). Nevertheless, the groups of 55 and 65+ years constituted 67.7% of the sample (n=107). The comparison between males and females is shown in Table 1. No significant differences were observed between groups in terms of age.

**Table 1.** Characteristics stratified by gender of individuals who died of COVID-19.

|  | All | Male | Female | X<sup>2</sup>, p |
| --- | --- | --- | --- | --- |
| Total | 158 | 95 (60.1) | 63 (39.9) |  |
| Age |  |  |  | 4.36, 0.35 |
| 24-34 | 5 (3.2) | 3 (3.2) | 2 (3.2) |  |
| 35-44 | 13 (8.2) | 9 (9.5) | 4 (6.3) |  |
| 44-54 | 33 (20.9) | 24 (25.3) | 9 (14.3) |  |
| 55-64 | 49 (31.0) | 29 (30.5) | 20 (31.7) |  |
| 65+ | 58 (36.7) | 30 (31.6) | 28 (44.4) |  |
| Any comorbidity |  |  |  | 2.55, 0.11 |
| Yes | 101 (63.9) | 56 (58.9) | 45 (71.4) |  |
| No or not reported | 57 (36.1) | 39 (41.1) | 18 (28.6) |  |
| Type 2 Diabetes-Yes | 56 (35.4) | 27 (27.4) | 29 (46.0) | 1.83, 0.17 |
| Obesity-Yes | 41 (25.9) | 21 (22.1) | 20 (31.7) | 5.13, 0.12 |
| Hypertension-Yes | 56 (35.4) | 31 (32.6) | 25 (39.7) | 0.82, 0.36 |

## Symptoms

The most common symptoms on admission were shortness of breath (overall: 88.77%; males: 55.2%, females: 44.8%), fever (overall: 71.42%, males: 51.4%, females: 48.6%), dry cough (overall: 64.28%: males: 58.7%, females: 41.3%), headache (overall: 43.87%, males: 55.8%, females: 44.2%), and myalgia (overall: 34.69%, males: 44.1%, females: 55.9%). When overlapping symptoms were observed, the first cluster included fever, dry cough, shortness of breath and headache. Other cluster was fever, dry cough, shortness of breath, myalgia, arthralgia and headache. Then fever, dry cough, shortness of breath, myalgia and arthralgia without headache. And fever and shortness of breath (Fig 2).

**Fig 2.** Venn diagram of overlapping symptoms. We plotted overlaps between the three main symptoms found in the sample. The higher proportions were fever or cough only, without manifestation of another symptom. The largest overlap was the triad fever-cough-dyspnea. Then, in order of frequency the main overlaps were fever-dyspnea and cough-dyspnea.

## Comorbidities

The most common comorbidities on admission were Type 2 Diabetes (60.63%), hypertension (59.57%) and obesity (43.61%). The main overlap between these comorbidities were T2D-Hypertensión. The second cluster was T2DM-Hypertension-Obesity and finally Hypertension-Obesity (Fig 3).

**Fig 3.** Venn diagram of overlapping comorbidities. The main overlap was the dyad Hypertension-T2D. The frequency of these comorbidities alone was almost equal. The next overlaps in order of frequency were the triad Hypertension-T2D-Obesity and the dyad Hypertension-Obesity.

## Pharmacological therapy

The most frequent drugs used for treating individuals with COVID-19 were azithromycin (60.6%) and hydroxychloroquine (53%). Less frequently employed were oseltamivir (27.3%), tocilizumab (16.77%) and lopinavir-ritonavir (16.7%). Drugs for various purposes (e.g. paracetamol, dexamethasone) were grouped as others, these were given to 66.7% of the patients.

Clustering drugs. We observed that azithromycin and hydroxychloroquine were frequently given together, followed by lopinavir-ritonavir with tocilizumab, and oseltamivir with lopinavir-ritonavir or tocilizumab (Fig 4).

**Fig 4.** Dendrogram of relationships between drugs used. The closest relation was azithromycin – hydroxychloroquine, followed by oseltamivir – lopinavir or tocilizumab.

## Discussion

In this study, we gathered clinical and demographic characteristics of individuals who died of COVID-19; we also identified drugs that were most used as antiviral treatment in those individuals.

There were 60% of male individuals. Gender consistency was an expected statistic as COVID-19 shows male predominance in Mexico’s epidemiological data (8), as well as in other populations (5, 14-16). We found that 67.7% of individuals were ≥55 years old. Although age varies widely among reports, our results are comparable to samples from patients with severe COVID-19 (14, 17). The mean length time hospitalized in days was 4.75 ± 4.43, which is shorter than what was reported in an observational study of fatal cases from Wuhan, China (6.35 ± 4.51) (17).

Shortness of breath, fever and dry cough were the most prevalent symptoms in our sample. Fever and cough had been reported as the most prevalent symptoms in several studies, including the Mexican population and severe cases (5, 10, 15, 17-19). On the other hand, the third most prevalent symptom varies widely; for instance, a review found dyspnea prevalence of 53 – 80%, and a series of fatal cases found in 70.6% (5, 17). Nonetheless, in clinical series and meta-analysis involving non-lethal cases, the third most prevalent symptom was myalgias or fatigue (31-67%) (10, 15, 18, 19). In our sample, shortness of breath was the most prevalent symptom and it was present in every cluster of symptoms, which could indicate that shortness of breath is an early symptom and bad prognosis.

The most common comorbidities in our sample were type 2 diabetes, hypertension and obesity. Remarkably, diabetes and hypertension had similar frequencies. In Mexican individuals with COVID-19, it is reported a slightly higher frequency of hypertension compared with diabetes (8, 10). Nonetheless, in other populations hypertension has a much higher prevalence compared to diabetes (5, 18). In two meta-analyses from the same work group (20, 21), hypertension and diabetes were associated with poor outcomes in COVID-19. The hypertension association was influenced by gender, but not by age or diabetes, while the diabetes association was influenced by age and hypertension. On the other hand, in a study using OpenSAFELY database (16), hypertension was associated with an increased risk (HR 1.09) but it could not be associated when the risk was adjusted to age, gender, diabetes and obesity. In addition, diabetes and obesity were associated with an increased risk of death (HR 1.90 and 1.92, respectively). Furthermore, epidemiological data of COVID-19 in Mexico, indicates that obesity is a risk factor for death and is a partial mediator on the effects of diabetes in decreased survival (8). Finally, when comorbidities in our sample were clustered, we observed a lower frequency of individuals with only-hypertension compared with only-diabetes and only-obesity patients. Therefore, diabetes and obesity should not be overlooked as strong risk factors and poor outcomes.

In our sample, the most frequent treatment was hydroxychloroquine associated with azithromycin. Although there is evidence of in vitro activity against SARS-CoV-2, the use of hydroxychloroquine and azithromycin for clinical benefit is supported by limited and conflicting clinical data (22). Antiviral agents were next in frequency order. Oseltamivir was the third most administrated treatment. However, there are no reports about oseltamivir having in vitro activity against SARS-CoV-2. Although some clinical trials include oseltamivir, it is not proposed as therapeutic intervention (23). Lopinavir-ritonavir was less frequently used as treatment in our sample; while it is reported in vitro activity against SARS-CoV-2, clinical trials have failed in associate it with better clinical outcomes (24). Finally, the only immunomodulatory drug used in our sample was tocilizumab. In a meta-analysis, tocilizumab was associated with a significant decrease in mortality rate and ameliorate clinical symptoms in patients with COVID-19 (25). Despite of its promising results, immunomodulatory drugs access is limited and it is not broadly used in clinical settings in Mexico.

Our study has some limitations. First, we had limited information about some individuals because they died within minutes after their arrival to the emergency room. Second, we did not include surviving individuals so we could not compare characteristics between groups. Third, we included data of only one frontline hospital for treating COVID-19. Consequently, this report shows characteristics and handling of patients at the beginning of the COVID-19 pandemic at the south of Mexico; therefore, our data cannot be applied to the whole Mexican population.

In conclusion, our study showed a high frequency of off-label use of drugs that have insufficient evidence of their efficacy against COVID-19. The individuals in our sample had shorter hospital stays, higher frequency of shortness of breath, and higher prevalence of diabetes compared with similar populations from other countries. It is important to do more clinical series and cohorts in Mexican population, as this population shows features that could differ of populations of other countries.

## Data Availability

any data can be requested directly by e-mail with the corresponding author

